# The impact of Gam-COVID-Vac, an AdV5/AdV26 COVID-19 vaccine, on the biomarkers of endothelial function, coagulation and platelet activation

**DOI:** 10.1101/2023.08.22.23294400

**Authors:** Anar Turmukhambetova, Sergey Yegorov, Ilya Korshukov, Valentina Barkhanskaya, Svetlana Kolesnichenko, Dmitriy Klyuyev, Zhibek Zhumadilova, Aruzhan Pralieva, Laylim Absaghit, Ruslan Belyaev, Dmitriy Babenko, Gonzalo H. Hortelano, Matthew S. Miller, Dmitriy Vazenmiller, Irina Kadyrova

## Abstract

**Background:** COVID-19 vaccines have played a critical role in controlling the COVID-19 pandemic. Although overall considered safe, COVID-19 vaccination has been associated with rare but severe thrombotic events, occurring mainly in the context of adenoviral vectored vaccines. A better understanding of mechanisms underlying vaccine-induced hypercoagulability and prothrombotic state is needed to improve vaccine safety profile.

**Methods:** We assessed changes to the biomarkers of endothelial function (endothelin, ET-1), coagulation (thrombomodulin, THBD and plasminogen activator inhibitor, PAI) and platelet activation (platelet activating factor, PAF, and platelet factor 4 IgG antibody, PF4 IgG) within a three-week period after the first (prime) and second (boost) doses of Gam-Covid-Vac, an AdV5/AdV26-vectored COVID-19 vaccine. Blood plasma collected from vaccinees (n=58) was assayed using ELISA assays. Participants were stratified by prior COVID-19 exposure based on their baseline SARS-CoV-2-specific serology results.

**Results:** We observed a significant post-prime increase in circulating ET-1, with levels sustained after the boost dose compared to baseline. ET-1 elevation following dose 2 was most pronounced in vaccinees without prior COVID-19 exposure. Prior COVID-19 was also associated with a mild increase in post-dose 1 PAI.

**Conclusions:** Vaccination was associated with elevated ET-1 up to day 21 after the second vaccine dose, while no marked alterations to other biomarkers, including PF4 IgG, were seen. A role of persistent endothelial activation followingCOVID-19 vaccination warrants further investigation.

## Introduction

COVID-19 vaccination has been associated with rare, but life-threatening, venous and arterial thrombotic events, with and without thrombocytopenia [1, 2]. Most reported thrombosis cases have been associated with the widely used adenoviral vectored vaccines, such as the ChAdOx1 CoV-19 vaccine (AstraZeneca, the University of Oxford) and the Ad26.COV2.S vaccine (Janssen; Johnson & Johnson) [1,2]. More recently, a fatal case of vaccine-induced immune thrombocytopenia and thrombosis (VITT) was reported after the receipt of the Gam-COVID-Vac (“Sputnik-V”, manufactured by the Gamaleya Research Institute of Epidemiology and Microbiology) vaccine in a 24-year-old woman, with the onset of symptoms on day 7 post-vaccination [3].

Although several haematological biomarkers have been linked to the VITT syndrome, including platelet number, d-dimer, fibrinogen, and platelet factor 4 (PF4)-specific immunoglobulin G (IgG), the exact mechanisms underlying vaccine-induced hypercoagulability and prothrombotic state remain enigmatic [1,2]. Recent studies conducted predominantly in Europe in general vaccinee cohorts have identified changes to clotting activation and endothelial function associated with the transient pro-inflammatory post-vaccination state[4-7]. Further investigation of these changes in the context of less well-studied vaccines such as the Gam-COVID-Vac, accounting for vaccination timing and prior natural exposure to SARS-CoV-2, is vital for refining vaccine strategies and enhancing their safety and efficacy globally. The Gam-COVID-Vac vaccine is a recombinant adenovirus (rAd) vectored vaccine, typically administered in two doses containing rAd26 as a prime and rAd5 as a boost, with a 21-day interval between the doses. Since late 2020, the vaccine has been deployed in 71 countries across Latin America, Africa, and Asia, including Kazakhstan. Between February and September 2021, the mass COVID-19 vaccination campaign in Kazakhstan relied primarily of Gam-COVID-Vac, with >85% of vaccinated subjects having received Sputnik-V [8]. During this massive vaccine rollout, we conducted a prospective study of safety, reactogenicity and immunologic responses to Gam-COVID-Vac in a cohort with and without prior history of COVID-19 exposure [9].

Following our earlier studies of “Sputnik-V” [9], here, we explored the effects of the vaccine on endothelial function, coagulation, and platelet activation and thus assessed changes to circulating endothelin (ET-1), thrombomodulin (THBD), plasminogen activator inhibitor (PAI), platelet activating factor (PAF) and PF4 IgG at three time points before and at 21 days following the prime and boost doses.

## Materials and Methods

### Study setting

The current analysis is a follow-up to an earlier observational prospective study of safety and reactogenicity of Gam-COVID-Vac in Kazakhstan. Venous blood samples were collected on after fasting at the same time in the morning. Due to a limited sample availability, only a subset of samples from the earlier study was analyzed. Briefly, the main study was conducted in Central Kazakhstan, where ∼60% of the population had had confirmed exposure to SARS-CoV-2 by Spring 2021 [9-11]. Participants were recruited at the Karaganda Medical University in April-May 2021 (ClinicalTrials.gov #NCT04871841) and consisted of asymptomatic adults who had not previously received a COVID-19 vaccine; individuals with respiratory symptoms or a laboratory-confirmed COVID-19 diagnosis within two weeks prior to the study were excluded. Short questionnaires were administered to gather information on participants’ demographic background and recent history of COVID-19 exposure. At follow-up, participants were screened for respiratory symptoms and tested for COVID-19; participants with COVID-19 at follow-up were excluded. Gam-COVID-Vac administration (0.5 ml dose of vaccine injected into the deltoid muscle) followed the national guidelines and was conducted after sample collection. The vaccine consisted of two doses: the first dose contained rAd26, and the second dose contained rAd5. Each dose contained 1± 0.5 x10^11^ rAd particles.

Based on serologic spike (S) IgG and IgA results, “no prior COVID-19” was defined as negative for both S-IgG and -IgA (IgG-, IgA-). The “no prior COVID-19” group was defined as positive for either or both IgG and/or IgA (IgG+/-, IgA+/-) at baseline.

### Biomarker assays

All ELISA assays were performed on blood plasma using commercially available ELISA kits (Cloud Clone Corp., China) for ET-1 (#CEA482Hu), PAI-1 (#SEA532Hu), TMBM (#SEA529Ca), PF4 IgG (#AEK505Hu) and PAF (#CEA526Ge) following the manufacturer protocol. Paired vaccinee samples were assayed on the same ELISA plate to avoid the effects of inter-plate variability. Absorbance was measured at OD450 nm using the Evolis 100 ELISA reader (Bio-Rad).

### Statistical analyses

All analyses and graphing were performed in JASP 0.17.2.1 and Prism 9.5.1 software. We used the Wilcoxon Signed Rank Test to assess differences across the time points, while differences between the Prior and No Prior COVID-19 groups were assessed using Mann-Whitney U or Pearson’s Chi-squared tests. Since ET-1 and PAF assays use the competitive inhibition principle, the OD values for these biomarkers were inverted prior to analysis by subtraction from the highest measured OD within each assay. Other assay OD values were analyzed without any transformation.

## Results

We analyzed a total of 58 plasma samples paired across three study visits (baseline, post-dose 1 and post-dose 2) that were available from the original clinical trial. The serologically confirmed presence of S-IgG and/or S-IgA was used to stratify the participants by prior COVID-19 as previously.

There were no demographic or biometric differences between the Prior/No Prior COVID-19 subgroups within the current study subset (Table 1). Due to the limited and variable sample availability, fewer samples were available for some of the assays, such as for PAF.

**Table 1.**
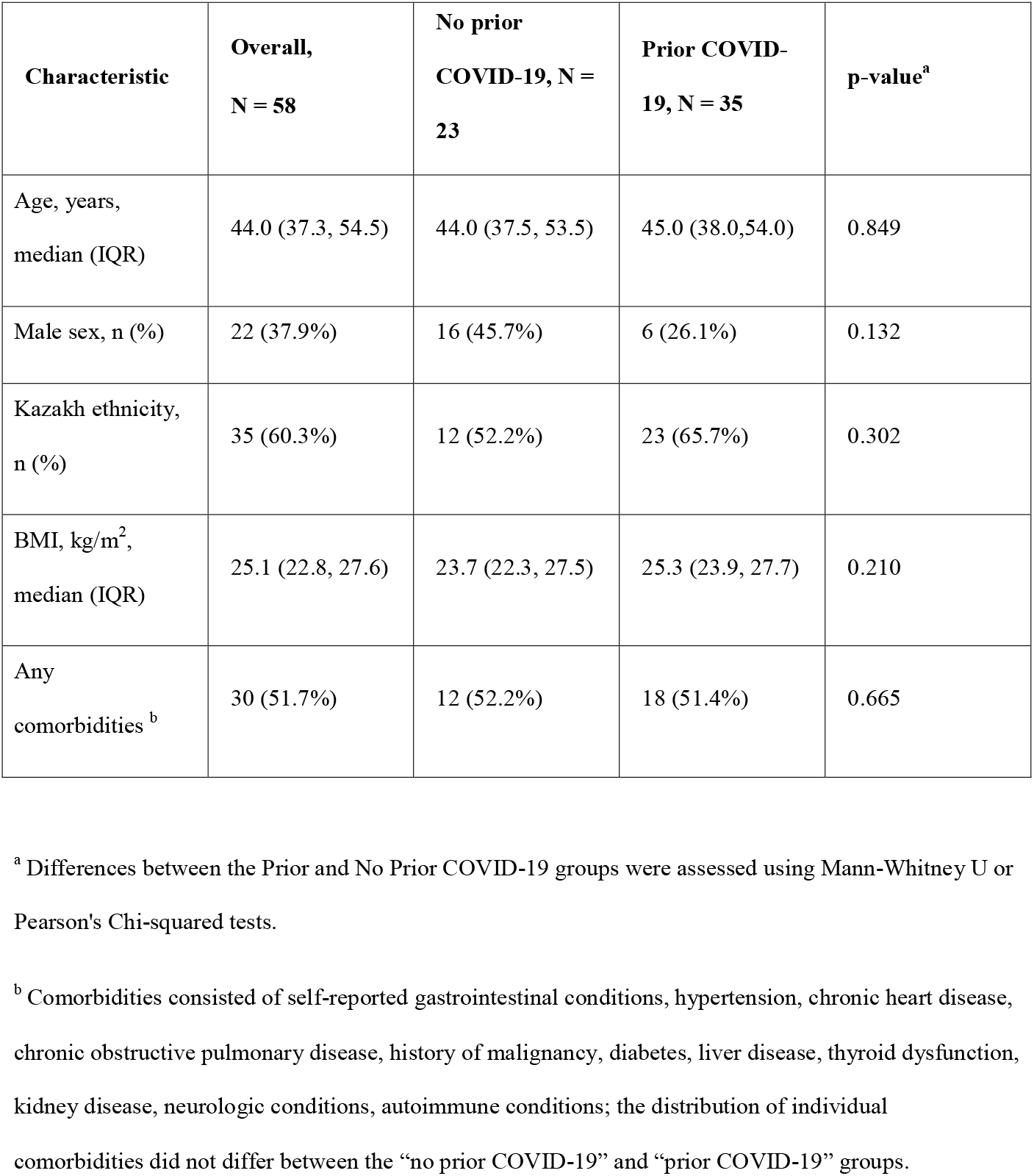
Characteristics of study participants.

In the main analysis (Fig 1), unstratified by prior COVID-19 exposure, ET-1 was the only marker significantly impacted by vaccination. ET-1 was significantly increased post-dose 1 (p=0.002), and this increase was sustained post-dose 2 compared to baseline (p=0.013).

**Fig 1.**
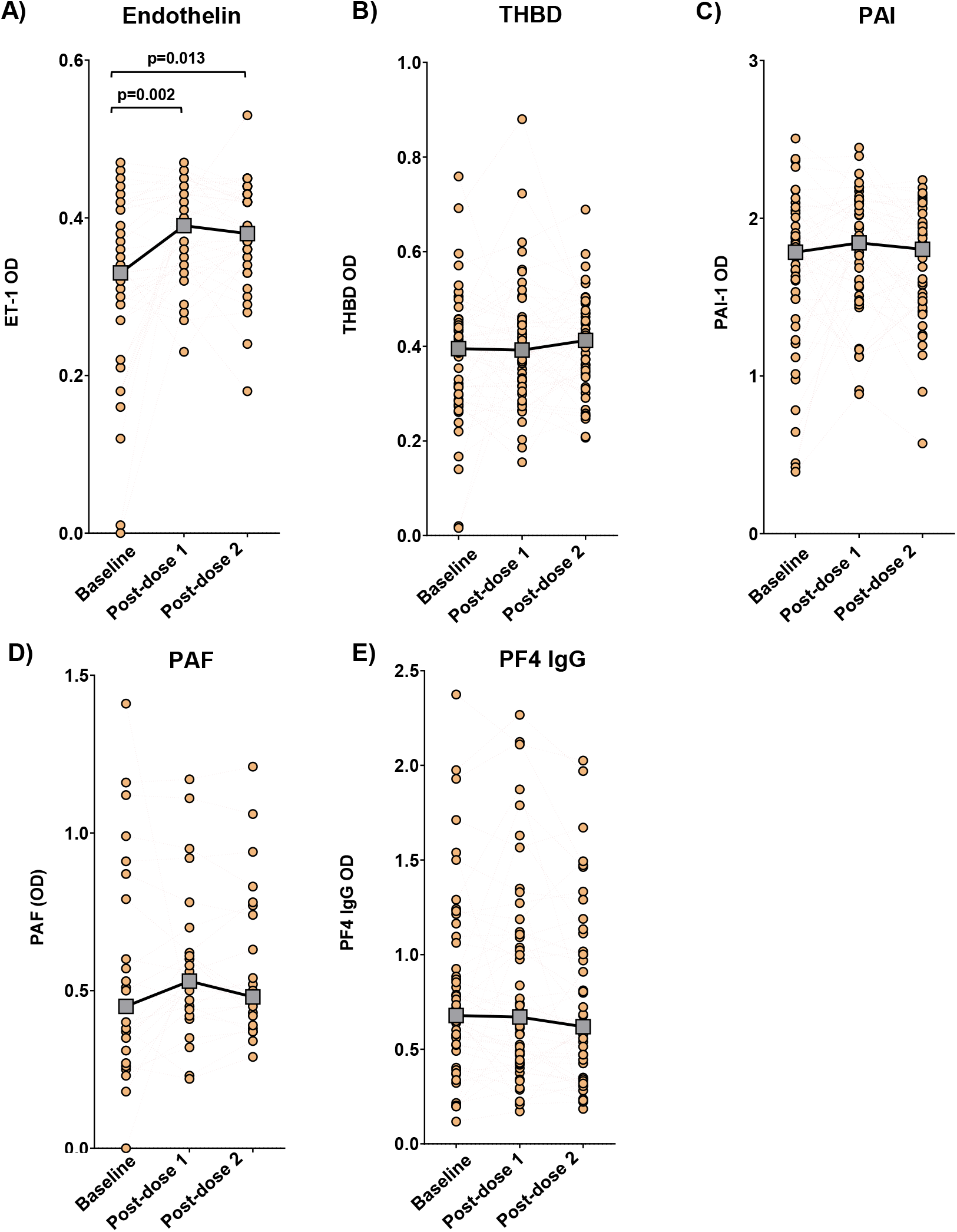
Impact of GamCovidVac vaccination on the blood biomarkers in all participants. A) Endothelin, ET-1 (n=45). B) Thrombomodulin, THBD (n=51). C) Plasminogen activator inhibitor, PAI (n=49). D) Platelet activating factor, PAF (n=27). E) Platelet factor 4 IgG antibody, PF4 IgG (n=50). Each dot denotes a study participant, dashed lines link samples paired across the study visits. Gray boxes denote median OD at each visit. OD: optic density. Statistical significance was assessed by the Wilcoxon Signed Rank Test.

In the analysis stratified by prior COVID-19 exposure (Fig 2), ET-1 was elevated post-vaccination in both prior and no prior COVID-19 groups, although this increase was not sustained in the prior COVID-19 group post-dose 2.

**Fig 2.**
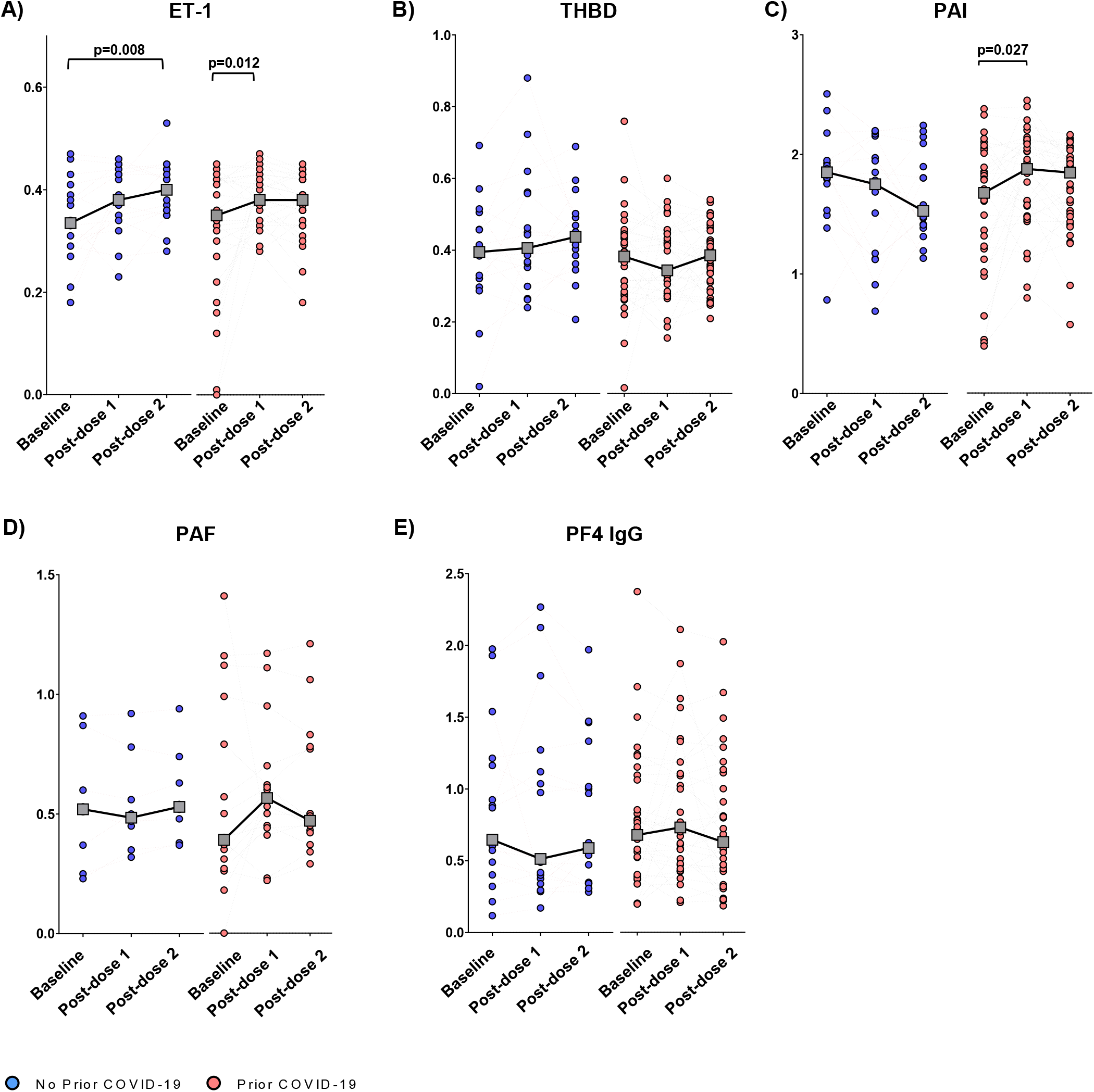
Impact of GamCovidVac vaccination on the blood biomarkers stratified by prior exposure to COVID-19. A) Endothelin, ET-1 (n=16 and 29). B) Thrombomodulin, THBD (n=17 and 34). C) Plasminogen activator inhibitor, PAI (n=16 and 33). D) Platelet activating factor, PAF (n=8 and 9). E) Platelet factor 4 IgG antibody, PF4 IgG (n=17 and 33). Each dot denotes a study participant, dashed lines link samples paired across the study visits. Gray boxes denote median OD at each visit. OD: optic density. Statistical significance was assessed by the Wilcoxon Signed Rank Test.

Lastly, a mild post-dose 1 increase (p=0.027) was observed for PAI only in the prior COVID-19 group.

## Discussion

Here we studied the impact of Gam-COVID-Vac on endothelial function, coagulation, and platelet activation biomarkers within a three-week period after the first and second vaccine doses. Vaccination significantly increased ET-1 levels after the first dose, and this elevation was sustained after the second dose. Furthermore, post-dose 2 ET-1 elevation was most pronounced in vaccinees without prior COVID-19 exposure. Prior COVID-19 was also associated with a mild increase in post-dose 1 PAI. Consistent with earlier studies [4,12], no marked alterations to other biomarkers, including PF4 IgG, were seen. To the best of our knowledge, our study is the first to assess thrombosis-associated biomarkers in the context of Gam-COVID-Vac vaccination. The current lack of studies on this aspect of “Sputnik-V” is partly explained by a shortage of data on the vaccine’s safety and performance outside of Russia, where the vaccine originated [13]. At the same time, the reported cases of VITT and myocarditis after the receipt of Gam-Covid-Vac have raised concerns about the under-reporting of vaccine-associated vascular events [3,14]. Our current findings support the data from both human and animal studies [5,13,15] that replication-deficient adenovirus vectored vaccines can trigger endothelial activation, which could potentially induce platelet aggregation and thrombosis.

ET-1, a potent vasoconstrictor produced by the vascular endothelium, is a key regulator of vascular tone and has been implicated in several cardiovascular diseases and in the pathogenesis of severe COVID-19 [16,17]. An increase in ET-1 level may therefore suggest a disturbance in endothelial function following vaccination. In support of this, studies of ChAdOx-1 have found elevated post-vaccination levels of Von Willebrand Factor, another biomarker of endothelial function [5].

Interestingly, when stratifying the data by prior COVID-19 exposure, we noticed a mild increase in PAI levels in the prior COVID-19 group following the first vaccine dose. PAI is a primary inhibitor of fibrinolysis and higher levels may hint towards an increased coagulation state, which is also observed in COVID-19 [2]. This finding suggests that individuals with prior COVID-19 exposure might exhibit a more pronounced coagulation response after the first vaccine dose, possibly due to a more robust immune response triggered by the recognition of the SARS-CoV-2 S protein in the vaccine.

Our study has several limitations, including the limited and variable sample availability, that precluded us from assaying more biomarkers. Our study cohort was also small and restricted to adults from Central Kazakhstan, limiting the generalizability of our findings. The observed changes to ET-1 and PAI, while statistically significant, were mild and their clinical significance is yet to be fully understood. Future studies with larger sample sizes and a more diverse population are needed to validate these findings and elucidate the clinical implications. Additionally, studies exploring other potential factors such as age, sex, comorbidities, and genetic predispositions that might influence these biomarker changes are warranted. Despite the limitations, our study provides valuable preliminary data on the changes in the biomarkers of endothelial function, coagulation, and platelet activation biomarkers associated with Gam-COVID-Vac vaccination. While the beneficial impact of COVID-19 vaccines in controlling the pandemic is undeniable, their association with a rare but severe prothrombotic state warrants further exploration, paying a particular attention to the role of vaccine-induced persistent endothelial activation.

## Supporting information

Table 1. Characteristics of study participants.

## Data Availability

All data produced in the present study are available upon reasonable request to the authors

## Declarations

### Ethics approval and consent to participate

All study procedures were approved by the Research Ethics Board of Karaganda Medical University under Protocol #18 from 12.04.2021. Written informed consent was obtained from all participants.

### Consent for publication

Authors provide consent for the publication of the manuscript detailed above, including any accompanying images or data contained within the manuscript.

### Availability of data and materials

The source data for all analyses will be made available with the article. Any additional data from this study will be made available, wherever possible, on appropriate request to the corresponding author.

### Authors contributions

AT: Conceptualization, Funding acquisition, Project administration, Resources, Supervision, Writing-original draft, Writing - review and editing. SY: Conceptualization, Data curation, Formal analysis, Investigation, Methodology, Supervision, Validation, Visualization, Writing-original draft, Writing - review and editing. IlK: Data curation, Formal analysis, Investigation, Writing-original draft, Writing - review and editing. VB: Investigation. SK: Formal analysis, Investigation. DK: Funding acquisition, Project administration, Resources, Supervision, Writing-original draft, Writing - review and editing. ZhZh: Investigation, Resources. AP: Investigation. LA: Investigation. RB: Investigation, Resources. GH: Conceptualization, Formal analysis, Investigation, Writing-original draft, Writing - review and editing. DB: Conceptualization, Data curation, Formal analysis, Investigation, Methodology, Visualization, Writing-original draft, Writing - review and editing. MSM-Conceptualization, Methodology, Supervision, Validation, Visualization, Writing - review and editing. DV: Conceptualization, Writing-original draft, Writing - review and editing. IK: Conceptualization, Data curation, Formal analysis, Funding acquisition, Investigation, Methodology, Project administration, Resources, Supervision, Visualization, Writing-original draft, Writing - review and editing. All authors had full access to all the data in the study and the lead authors (SY, IK, DV) had final responsibility for the decision to submit manuscript for publication.

All authors read and approved the final manuscript.

## Acknowledgements

We thank all the participants and clinical and laboratory staff, who have been involved in the study.

## Supplementary information

All supplementary information can be found in the Appendix.

