## Supplementary material for "The impact of Gam-COVID-Vac, an AdV5/AdV26 COVID-19 vaccine, on the biomarkers of endothelial function, coagulation and platelet activation": Table 1. Characteristics of study participants.

| **Characteristic** | **Overall,**  **N = 58** | **No prior COVID-19, N = 23** | **Prior COVID-19, N = 35** | **p-value^*^** |
| --- | --- | --- | --- | --- |
| Age, years, median (IQR) | 44.0 (37.3, 54.5) | 44.0 (37.5, 53.5) | 45.0 (38.0,54.0) | 0.849 |
| Male sex, n (%) | 22 (37.9%) | 16 (45.7%) | 6 (26.1%) | 0.132 |
| Kazakh ethnicity, n (%) | 35 (60.3%) | 12 (52.2%) | 23 (65.7%) | 0.302 |
| BMI, kg/m^2^, median (IQR) | 25.1 (22.8, 27.6) | 23.7 (22.3, 27.5) | 25.3 (23.9, 27.7) | 0.210 |
| Any comorbidities ^ | 30 (51.7%) | 12 (52.2%) | 18 (51.4%) | 0.665 |

*Differences between the Prior and No Prior COVID-19 groups were assessed using Mann-Whitney U or Pearson's Chi-squared tests.

^ Comorbidities consisted of self-reported gastrointestinal conditions, hypertension, chronic heart disease, chronic obstructive pulmonary disease, history of malignancy, diabetes, liver disease, thyroid dysfunction, kidney disease, neurologic conditions, autoimmune conditions; the distribution of individual comorbidities did not differ between the “no prior COVID-19” and “prior COVID-19” groups.
